# Peripheral Immune pattern in a genetic cohort of p.A53T alpha-synuclein Parkinson’s disease

**DOI:** 10.1101/2024.11.14.24317039

**Authors:** Christos Koros, Athina-Maria Simitsi, Nikolaos Papagiannakis, Roubina Antonelou, Anastasia Bougea, Dimitra Papadimitriou, Ioanna Pachi, Ion Beratis, Dionysia Kontaxopoulou, Stella Fragkiadaki, Evangelos Sfikas, Ioanna Alefanti, Chrysa Chrysovitsanou, Efthalia Angelopoulou, Marianna Bregianni, Konstantinos Lourentzos, Vasilios C. Constantinides, Georgios Velonakis, Vasilios Prassopoulos, Anastasios Bonakis, Constantin Potagas, Sokratis G. Papageorgiou, Maria Stamelou, Leonidas Stefanis

**Author notes:** Corresponding Author: Christos Koros, MD, PhD, 1st Department of Neurology, Eginition Hospital, National and Kapodistrian University of Athens, Athens, Greece.

## Abstract

**Introduction:** Previous research has shown that inflammatory immune biomarkers including peripheral white blood cell subpopulations differ between Parkinson’s disease (PD) patients and healthy controls (HC), with idiopathic PD exhibiting higher neutrophil to lymphocyte ratio (NLR). The aim of our present report was to assess the peripheral immune profile in patients or asymptomatic carriers harboring the p.A53T alpha-synuclein (SNCA) mutation.

**Methods:** Data regarding 31 p.A53T SNCA PD patients, 9 asymptomatic mutation carriers and 194 HCs were obtained from the database of the Parkinson’s Progression Markers Initiative (PPMI). Focus was placed on peripheral immune blood cells subpopulations and clinical/imaging parameters during the initial study assessment.

**Results:** NLR, Absolute Neutrophil cell count and Neutrophil to total Leukocytes ratio were increased in the p.A53T SNCA PD group as compared to HCs [2,77 vs 2,18 (p<0.001), 4,32×10^3 cells/μL vs 3,67x 10^3 cells/μL (p=0.001), 65,67% vs 59,55% (p<0.001) respectively]. Differences in NLR were mainly driven by the male patient subgroup. The absolute Lymphocyte cell count showed a trend towards being decreased in p.A53T PD, and Lymphocyte to total leukocytes ratio was lower in p.A53T SNCA cohort as compared to HC [26,16% vs 30,02% (p=0.001)]. Monocyte to total Leukocytes ratio was lower in p.A53T PD 5,49% vs 6,74% (p=0.002). Finally, we observed a positive correlation between the absolute Lymphocyte count and the mean putamen DATSCAN signal. Asymptomatic carriers did not differ statistically from p.A53T SNCA PD or HC regarding leucocyte subpopulations counts.

**Discussion:** Our current study provides evidence of a specific pattern of peripheral immune response in the p.A53T SNCA PD group which aligns well with literature data in idiopathic and other genetic PD forms. Furthermore, given former evidence that alpha-synuclein represents an immune target in PD, we can speculate a putative underlying inflammatory pathway in this archetypal form of genetic synucleinopathy.

## Introduction

Parkinson’s disease (PD) represents the second most common neurodegenerative disorder. A number of biomarkers are used in clinical practice in order to monitor the progression of the disease. Such biomarkers include cerebrospinal fluid (CSF) alpha-synuclein and other biomarkers used in the evaluation of other forms of neurodegeneration like Alzheimer’s disease (CSF total tau protein and A-beta amyloid Aβ42 levels). Moreover other plasma or CSF biomarkers of neuronal and glial destruction or synaptic integrity have been applied (glial fibrillary acid protein (GFAP), serum neurofilament light chain (NFL) or small extracellular vesicle (sEV)) (Park et al., 2024, Hirama et al., 2024, Chopra and Outeiro, 2024).

Notably, except for the established importance of neurodegeneration, the role of neuroinflammation in PD is increasingly recognized (Williams et al., 2022). Recent studies have shown that inflammatory biomarkers are increased in the peripheral blood of PD patients (Akil et al., 2015). It appears that peripheral inflammatory conditions play a key role in the pathophysiology of PD and apart from cytokines, also peripheral immune cells are important (Dommershuijsen et al., 2022). A dysregulation of cytokines in the CSF, brain tissue and blood of PD patients has been observed. Inflammatory immune cells migrate from the periphery to the central nervous system (CNS) and enhance neuroinflammation. This procedure might trigger dopaminergic neuron cell death as it is the case in PD (Hosseini et al., 2023).

Previous studies have reported alterations in the subpopulations of leukocytes in peripheral blood of PD patents. The neutrophil-to-lymphocyte ratio (NLR) represents a widely used biomarker in many conditions ranging from intensive care unit patient assessment to neurodegenerative disorders and normal pressure hydrocephalus (Kara et al. 2022, Bissacco et al., 2024, Novellino et al., 2021). Its role as an emerging biomarker in Alzheimer’s disease (AD) is adequately supported by former research (Bawa et al., 2020). NLR is particularly relevant to the peripheral inflammation in PD (Muñoz-Delgado et al., 2021). A lower lymphocyte count is associated with increased risk of PD (Jensen et al., 2021). Other markers like lymphocyte-monocyte ratio (LMR), neutrophil-to-high-density lipoprotein ratio (NHR) and platelet-to-lymphocyte ratio (PLR) have been studied as inflammatory indicators in neurodegenerative disorders (Liu et al., 2021, Li et al., 2024). In rodent models of PD an infiltration by T-cells has been reported in various CNS areas, including the striatal perivascular space, neocortex and hippocampus. A higher lymphocyte ratio has been shown to be protective in terms of neurodegeneration. A number of studies have shown higher NLR in patients with PD and this marker could discriminate idiopathic PD from healthy controls.

Individuals with a higher lymphocyte count exhibit reduced vulnerability to the development of PD. However, there are controversial outcomes in the literature with some studies favoring a positive correlation between the NLR and disease duration while this was not evident in other reports. In contrast, neutrophils can have a pro-inflammatory action and enhance inflammatory response by means of chemokines. Furthermore, the integrity of the blood-brain barrier (BBB) is closely related to the plasma level of high-density lipoprotein cholesterol (which is lower in PD). Neutrophil-to-high-density lipoprotein ratio is increased in PD patients and shows a negative correlation with duration of the disorder (Li et al., 2024). Additionally, there is evidence that peripheral blood monocytes are also altered in PD and a putative proinflammatory impact of α-synuclein on monocytes have been suggested as a putative mechanism (Su et al., 2022). There is paucity in the literature regarding the application of these markers in REM Sleep Behavior Disorder (RBD) but LMR and CRP level could play a role in evaluating patient prognosis in PD combined with RBD (Wang et al., 2022).

Regarding the effect of these markers to the clinical course of PD there are many recent research data supporting a relationship (Grillo et al., 2023, Kim et al., 2024, Umehara et al., 2020). The plasma NLR and NHR have been shown to be higher in patients with PD than in healthy controls, while the plasma LMR was substantially lower. The plasma NLR and NHR were positively correlated with Unified Parkinson’s Disease Rating Scale (UPDRS) (and subscores including UPDRS-I, UPDRS-II, and UPDRS-III) and Hoehn and Yahr staging scale (H&Y). There was a negative relationship of NLR with Mini-Mental State Examination (MMSE) and Montreal Cognitive Assessment (MoCA) scores (Contaldi et al., 2022), suggesting again that NLR is positively correlated with disease severity, in this case cognitive dysfunction.

As far as imaging data are concerned, Zhang and co-authors showed that higher NLR was associated with lower DAT levels in the caudate and the putamen. The lymphocyte count correlated positively with striatal DAT levels. The authors conclude that dopaminergic degeneration is associated with peripheral inflammation in PD (Zhang et al., 2024). It appears thus that the NLR, a widely used inflammatory marker, may have the potential to reflect the degree of dopaminergic degeneration in individuals with early-stage PD (Muñoz-Delgado et al., 2023a).

Apart from idiopathic PD, a limited number of previous studies have assessed the impact of genetic forms of PD (GBA1 and LRRK2 mutation carriers) on peripheral immune parkers including leukocyte subpopulations (Muñoz-Delgado et al., 2023b). Given former data supporting the importance of peripheral leukocyte subpopulations in idiopathic PD and other forms of synucleinopathies, the aim of our present report was to assess the peripheral immune profile in a genetic cohort of PD patients harboring the p.A53T alpha-synuclein (SNCA) mutation and its clinical impact.

## Methods

Data regarding 31 p.A53T SNCA PD patients, 9 asymptomatic p.A53T SNCA carriers and 194 HCs were obtained from the database of the Parkinson’s Progression Markers Initiative (PPMI). Focus was placed on peripheral immune blood cells subpopulations, clinical data including MDS-UPDRS part III in OFF and ON, MoCA score and DATSCAN imaging signal measurements during the initial study assessment.

Data used in the preparation of this article were obtained on 03/27/2023 (08/15/2024 last update) from the PPMI database (http://www.ppmiinfo.org/access-data-specimens/download-data),RRID:SCR 006431. For up-to-date information on the study, visit http://www.ppmi-info.org. The present study was conducted in agreement with the principles of the Declaration of Helsinki. Signed informed consent was obtained from all participants recruited. The study was approved by the Scientific Board of all PPMI sites involved (including the Scientific Board of Eginition hospital).

Hematological and Biochemical analyses (including measurements of peripheral blood leukocytes subpopulations) have been carried out in Covance laboratories in a uniform fashion, as per the study protocol.

Statistical analysis for baseline comparisons between the p.A53T SNCA PD subjects vs Healthy controls (HC) and asymptomatic carriers vs SNCA PD or HC was performed using univariate Analysis of Covariance (ANCOVA). Factors that could have an impact on measurements including age and sex were used as covariates in the analysis. Statistical significance was set at p<0.05.

Pearson correlations were calculated between NLR, neutrophils or leukocytes counts and various baseline parameters (basal ganglia subregions DATSCAN signal measurements, MDS-UPDRS score part III in OFF and ON and Montreal Cognitive Assessment (MoCA) score) in the p.A53T SNCA PD cohort. Statistical significance was set at p<0.05.

## Results

Demographic and clinical data regarding the p.A53T SNCA PD and asymptomatic carriers cohort have been included in Table 1. The mean age for PD was 50,58±10,48 years and the mean disease duration was 3,81±2,73 years. The average MDS-UPDRS III score in OFF was 23,21±16,35 while in ON was 18,63±12,19. The mean MoCA score was 25,03±5,28. The mean age for asymptomatic was 43,44±13,49 years the mean MoCA score was 27,11±2,21. (Table 1).

**Table 1.** Epidemiological and clinical features of the p.A53T SNCA PD and asymptomatic cohorts. (Mean ± SD)

| Cohort Features | p.A53T SNCA PD (N=31) | Asymptomatic p.A53T SNCA (N=9) |
| --- | --- | --- |
| Age (years) | 50,58±10,48 | 43,44±13,49 |
| Sex | 16 F / 15 M | 8 F / 1 M |
| Disease Duration (years) | 3,81±2,73 | N/A |
| MDS-UPDRS III score in "OFF" | 23,21±16,35 | 0,11±0,33 |
| MDS-UPDRS III score in "ON" | 18,63±12,19 | N/A |
| H & Y | 1,8±0,71 | 0 |
| MoCA score | 25,03±5,28 | 27,11±2,21 |
| DATSCAN Caudate Right | 1,29±0,53 | N/A |
| DATSCAN Caudate Left | 1,26±0,35 | N/A |
| DATSCAN Caudate Means | 1,28±0,44 | N/A |
| DATSCAN Putamen Right | 0,60±0,30 | N/A |
| DATSCAN Putamen Left | 0,64±0,41 | N/A |
| DATSCAN Putamen Means | 0,62±0,36 | N/A |
| White Blood Cells Absolute Number | 6,50±1,82 $\times 10^3$ | 5,77±1,10 $\times 10^3$ |
| Neutrophils Absolute Number | 4,32±1,45 $\times 10^3$ | 3,59±0,89 $\times 10^3$ |
| Lymphocytes Absolute Number | 1,66±0,54 $\times 10^3$ | 1,7±0,51 $\times 10^3$ |
| Monocytes Absolute Number | 0,36±0,15 $\times 10^3$ | 0,34±0,1 $\times 10^3$ |
| Eosinophils Absolute Number | 0,13±0,08 $\times 10^3$ | 0,1±0,05 $\times 10^3$ |
| Basophils Absolute Number | 0,04±0,02 $\times 10^3$ | 0,04±0,02 $\times 10^3$ |

In our study the NLR ratio was significantly increased in the p.A53T SNCA PD group as compared to HCs [2,77 vs 2,18 (p<0.001). Differences in NLR were mainly driven by the male patient subgroup [male p.A53T SNCA PD vs HC (p<0.001) as compared to female p.A53T SNCA PD vs HC (p=0.071)] (Figure 1). Neutrophil to Monocyte ratio (NMR) was also increased as compared to HCs 12,85 vs 9,52 (p<0.001).

**Figure 1.**
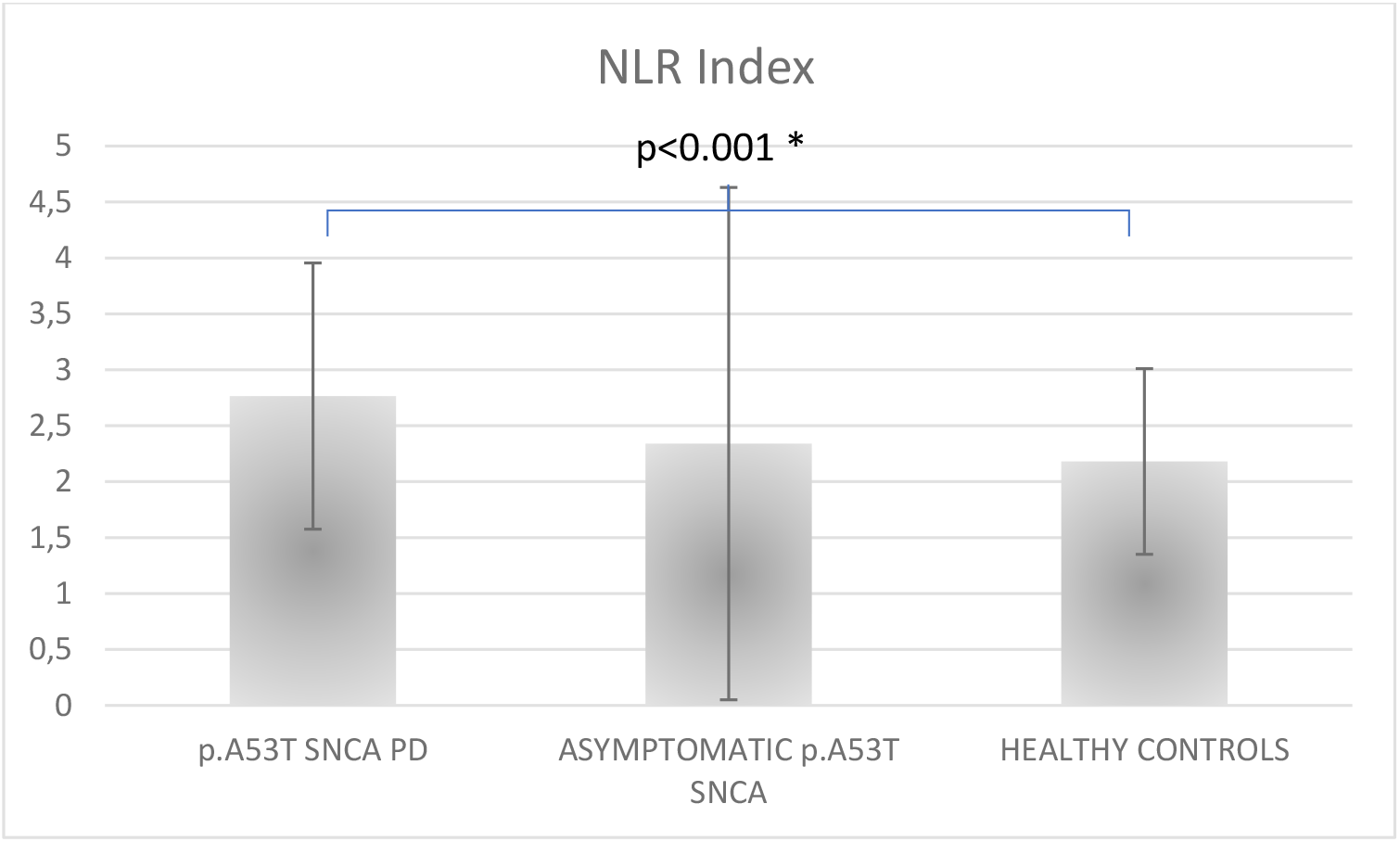
NLR index in p.A53T SNCA carriers (PD and asymptomatic) and healthy control group (Means±SD).

Moreover, the absolute Neutrophil cell count and Neutrophil to total Leukocytes ratio were also higher than in HC [4,32×10^3 cells/μL vs 3,67x 10^3 cells/μL (p=0.001), 65,67% vs 59,55% (p<0.001) respectively] (Figure 2). Differences in Neurtophil absolute cell count and ratio were again mainly driven by the male patient subgroup [male p.A53T SNCA PD vs HC (p<0.001 for absolute count and p<0,001 for ratio) as compared to female p.A53T SNCA PD vs HC (p=0.555 for absolute count and p=0.013 for ratio].

**Figure 2.**
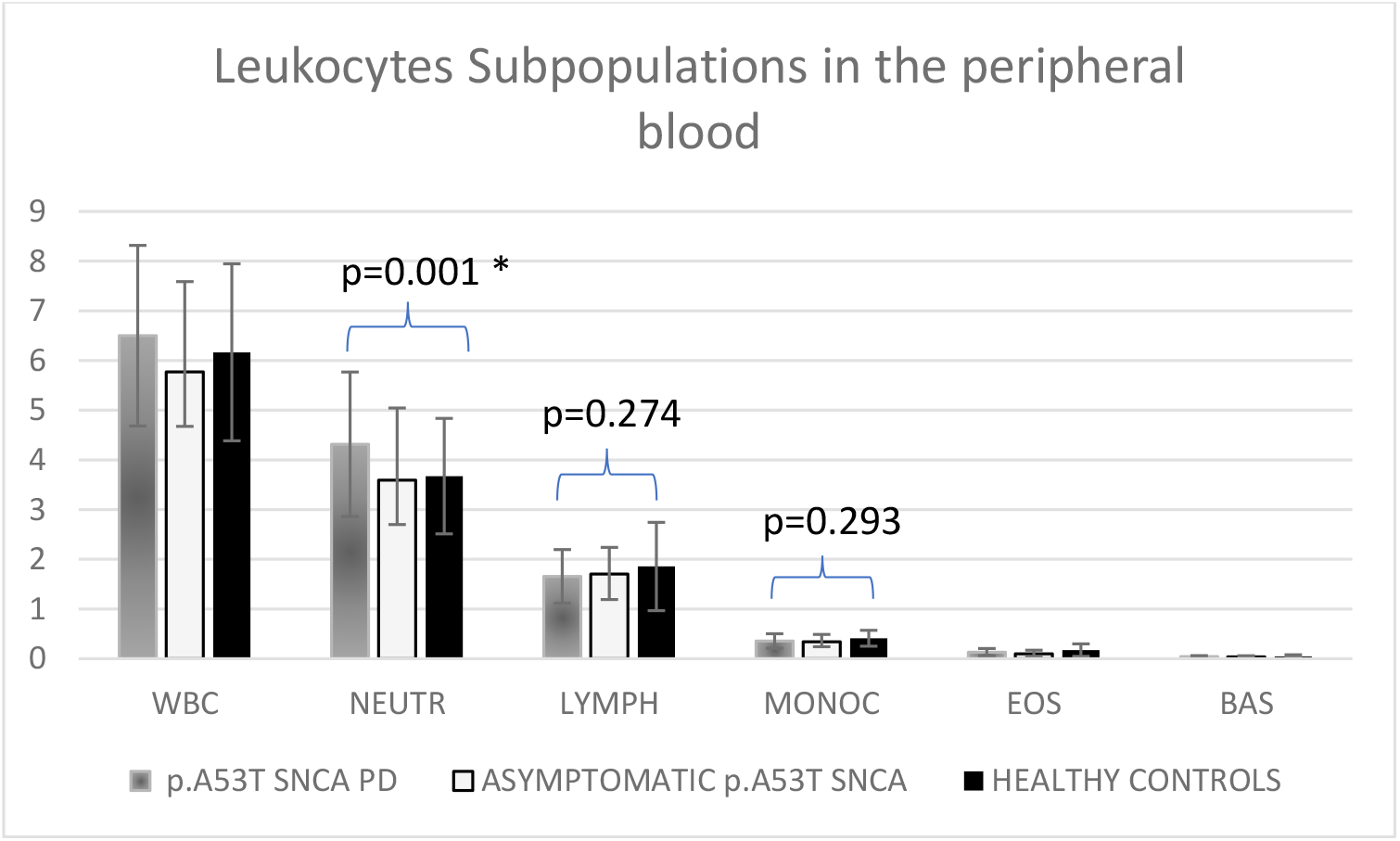
Peripheral Leukocyte subpopulations in p.A53T SNCA carriers (PD and asymptomatic) and healthy control group (Means±SD).

- WBC: White Blood Cells, NEUTR: Neutrophils, LYMPH: Lymphocytes, MONOC: Monocytes, EOS: Eosinophils, BAS: Basophils, SNCA: Alpha-Synuclein

The absolute Lymphocyte cell count showed a trend towards being decreased in p.A53T PD but did not reach significance [1,66 x10^3 cells/μL vs 1,86x 10^3 cells/μL (p=0,274)].

However, Lymphocyte to total leukocytes ratio was lower in p.A53T SNCA cohort as compared to HC [26,16% vs 30,02% (p=0.001)]. In contrast to NLR and Neutrophils, differences in Lympocyte absolute cell count and ratio were not sex specific [male p.A53T SNCA PD vs HC (p=0.93 for absolute count and p=0.013 for ratio) as compared to female p.A53T SNCA PD vs HC (p=0.045 for absolute count and p=0.027 for ratio].

Similarly, the absolute Monocyte cell count showed a trend towards being decreased in p.A53T PD but did not reach significance [0,36 x10^3 cells/μL vs x 0,41 x10^3 cells/μL (p=0,293)]. Monocyte to total Leukocytes ratio was lower in p.A53T PD 5,49% vs 6,74% (p=0.002).

Regarding less prominent subpopulations of blood leukocytes, the absolute Eosinophil cell count and Eosinophil to total Leukocyte ratio showed a trend towards being decreased in p.A53T PD but did not reach significance [0,13 x10^3 cells/μL vs x 0,17 x10^3 cells/μL (p=0,293) and 2,03% vs 2,84% (p=0,168) respectively].Finally, the absolute Basophil cell count and Basophil to total Leukocytes ratio showed a trend towards being decreased in p.A53T PD but did not reach significance 0,04 x10^3 cells/μL vs x 0,05×10^3 cells/μL

(p=0,551) and 0,68 % vs 0,81% (p=0.305) respectively].

We observed a positive correlation between the absolute Lymphocyte count and the mean putaminal DATSCAN signal [Pearson correlation: r = 0.631 (p = 0.005)]. There was no significant correlation between the absolute Neutrophil count or the absolute Lymphocyte count and MDS-UPDRS part III scores in OFF or ON, the MoCA score and mean caudate DATSCAN signal. Furthermore, despite a trend towards a positive correlation between NLR and MDS-UPDRS III in OFF [Pearson correlation: r = 0.343 (p = 0.064)], there was no statistically significant correlation between NLR and MDS-UPDRS part III scores in OFF or ON, the MoCA score and mean putaminal or caudate DATSCAN signal. Finally, we could not observe any correlation between disease duration in the p.A53T SNCA PD group and any of the indices of peripheral inflammation (NLR: Pearson correlation r = 0.166 (p = 0.373)].

As far as asymptomatic p.A53T SNCA carriers are concerned, no statistically significant differences between asymptomatic carriers and either p.A53T SNCA PD or HC could be evidenced regarding leucocyte subpopulations counts despite a trend for intermediate measurements between PD and HC [Asymptomatic carriers vs PD, NLR : 2,34 vs 2,77 (p=0,873), Absolute Neutrophil count: 3,59 vs 4,32 (p=0,903), Absolute Lymphocyte count: 1,70 vs 1,66 (p= 0,567) and Absolute Monocyte count: 0,34 vs 0,36 p= 0,825). Similarly, Asymptomatic carriers vs HC, NLR: 2,34 vs 2,18 (p=0,178), Absolute Neutrophil count: 3,59 vs 3,67 (p=0,730), Absolute Lymphocyte count: 1,70 vs 1,86 (p= 0,595) and Absolute Monocyte count: 0,34 vs 0,41 p= 0,774)].

## Discussion

The impact of peripheral immunity on genetic forms of PD still remains elusive. In the present study we have assessed peripheral blood subpopulation profile in a genetic cohort harboring the p.A53T mutation in the alpha-synuclein (SNCA) gene. The latter represents an archetypal form of genetic synucleinopathy with many features of idiopathic Parkinson’s disease being exacerbated. Patients carrying this pathogenic variant exhibit a unique and variable phenotype which resembles idiopathic PD, albeit with a more rapid motor and cognitive deterioration and occasionally pronounced non-motor symptoms like autonomic dysregulation and cognitive dysfunction (Papadimitriou et al., 2016, Koros, Stamelou et al., 2018).

Our study results provide evidence of a specific pattern of peripheral immune response in the p.A53T SNCA PD group (increased NLR ratio, increased neutrophils and reduced lymphocytes and monocytes) which is consistent with observations in idiopathic PD (Muñoz-Delgado et al., 2021). We have also observed a positive correlation between the absolute

Lymphocyte count and the mean putaminal DATSCAN signal in this genetic PD group while other correlations between NLR, absolute Neutrophil or Lymphocyte count and other clinical or imaging parameters were not significant. In a previous report, Jensen et al compared 465 idiopathic PD cases and 312,125 controls (Jensen et al., 2021). Lower lymphocyte count was associated with increased risk of subsequent PD diagnosis. There was some evidence that reductions in eosinophil counts, monocyte counts and CRP were associated with clinical PD and that higher neutrophil count was also associated. Only the association between lower lymphocyte count and PD remained robust to sensitivity analyses and further analysis suggested that the effect of lower lymphocyte count on PD may be causal. Immune parameters were correlated with CSF levels of total α-synuclein, amyloid-β-42, total and phosphorylated-tau and main motor and non-motor scores. This study provided in vivo evidence that, in PD, changes in leukocytes in the periphery, assessed as relative lymphopenia and NLR increase, reflect in central neurodegeneration-associated protein modifications, especially in α-synuclein and amyloid-β pathways, and greater clinical burden (Jensen et al., 2021).

In another study using data obtained from the Parkinson’s Progression Markers Initiative cohort as in our case, Kim and co-authors assessed whether peripheral blood neutrophils and lymphocytes are associated with longitudinal motor and cognitive decline in patients with early idiopathic PD (Kim et al., 2023). According to their findings patients with PD showed higher neutrophil and lower lymphocyte counts, resulting in a higher NLR than that in HC. Higher neutrophil counts were associated with a greater increase in MDS-UPDRS part 3 scores in patients with PD. Correspondingly, higher neutrophil levels were related to a greater reduction in DAT activity in the caudate. Among the blood biomarkers, only a higher neutrophil count was associated with faster motor progression along with accelerated nigrostriatal dopaminergic degeneration in patients with PD. The impact of neutrophils and lymphocytes on longitudinal cognitive changes remained unclear.

The study group of Munoz Delgado assessed clinical features, the peripheral immune profile, and striatal [123 I]FP-CIT DAT binding levels of 211 patients with PD (primary-cohort) and for replication purposes, they also studied a separate cohort of 344 de novo patients with PD enrolled in the Parkinson’s Progression Markers Initiative (PPMI-cohort) (Muñoz-Delgado et al., 2021, 2023a). A higher NLR was significantly associated with lower DAT levels in the caudate and the putamen. Lower lymphocyte count was significantly associated with lower DAT levels in both the caudate and the putamen but an association with the neutrophil count was not consistently observed. The accordance of outcomes across two independent cohorts suggests a relationship between systemic inflammation and dopaminergic degeneration in patients with PD, mainly driven by the lymphocyte count.

The same group in an additional study evaluated the impact of the genetic background of patients with PD on the peripheral inflammatory immune response (Muñoz-Delgado et al., 2023b). Patients with sporadic PD and Glucocerebrosidase gene (GBA)-associated PD showed a significantly lower lymphocyte count, a non-significantly higher neutrophil count and a significantly higher NLR than HC. The peripheral inflammatory immune response of patients with leucine-rich repeat kinase 2 (LRRK2)-associated PD did not differ from HC. This study supports the involvement of a peripheral inflammatory immune response in the pathophysiology of sporadic PD and GBA-associated PD. However, this inflammatory response was not found in LRRK2-associated PD, probably reflecting different pathogenic inflammatory mechanisms.

In accordance to the results of Munoz-Delgado et al related to genetic PPMI cohorts, our current study provides evidence of a specific pattern of peripheral immune response in the p.A53T SNCA PD group which aligns well with literature data in certain genetic PD forms (like GBA1 carriers). Different outcomes as compared to carriers of mutations in other PD genes like LRRK2 need to be further replicated and analyzed. Furthermore, given former evidence that alpha-synuclein represents an immune target in PD, we can speculate a putative exaggerated underlying inflammatory pathway in this archetypal form of genetic synucleinopathy (Lindestam Arlehamn et al., 2020).

PD patient populations carrying pathogenic mutations in recessive PD genes like PRKN and PINK1 have not been studied in terms of peripheral immune alterations with the exception of the Cooperative Health Research in South Tyrol (CHRIS) study which assessed heterozygous PRKN mutation carriers in a large population sample and reported a higher NLR ratio as compared to non carriers. In terms of clinical symptomatology, a higher number of heterozygous mutation carriers also manifested a detectable increase in an akinesia-related phenotype, diabetes and lower resting heart rate (Castelo Rueda et al., 2021).

Our study is one of a limited number of published reports, addressing the impact of genetic forms of PD on peripheral immunity background and the first to assess SNCA mutation carriers. An important merit of our current study is that data we used from the PPMI database have been collected and processed uniformly across PPMI centers, with a thorough standardized clinical and laboratory assessment. On the other hand, a major limitation is the relatively small number of A53T SNCA subjects, albeit a rare condition. Finally, it appears that the A53T SNCA group has a larger standard deviation (SD) concerning Leukocyte subpopulations counts levels than HC. This could be attributed to the relatively small size of these subgroups and/or to the heterogeneity in this population.

The pathogenic role of peripheral immunity on conversion of prodromal or asymptomatic individuals to PD remains obscure. There is a debate whether the observed abnormalities in the inflammatory response triggers dopaminergic neuron cell death and contributes to the disease manifestation or this is merely a response to the disease (reverse causation). One could speculate that a certain level of peripheral inflammation is needed for the disease to manifest especially for genetic forms, given the high but incomplete penetrance. This is consistent with data from idiopathic PD and genetic forms (SNCA and GBA1 mutation carriers but results regarding the LRRK2 PD carriers would argue against these possibilities). In general, there is accumulating evidence supporting the crucial involvement of T lymphocytes subpopulations in PD. In some previous studies an increased proportion of Th1 and Th17 cells (which have a pro-inflammatory effect) and a decreased number of Th2 cells (which exert anti-inflammatory functions) has been recorded in PD, leading to a pro-inflammatory Th1-biased immune response in these patients (Chen et al., 2018, Munoz-Delgado et al.,2023b). According to the aforementioned studies, an increased proportion of neutrophils has been linked to chronic inflammation whereas decreased populations of lymphocytes possibly represent an inadequate regulatory pathway. Moreover, another argument in favor of a pathogenic role of inflammatory dysregulation in PD is that in many previous studies, peripheral inflammation (as exemplified by markers like NLR) is robustly related to disease severity (Grillo et al., 2023, Kim et al., 2024, Umehara et al., 2020, Contaldi et al., 2022). In our current SNCA PD cohort, we observed a positive correlation between the absolute

Lymphocyte count and the mean putaminal DATSCAN signal which is consistent with a putative protective role of lymphocytes in dopaminergic denervation observed in PD. Finally, we have to underline the fact that the peripheral leukocyte profile of asymptomatic p.A53T SNCA carriers was either similar to HC or intermediate between HC and PD implying that leukocyte subpopulation disequilibrium might occur with the conversion to full-blown clinical PD.

Apart from PD, the aforementioned inflammatory biomarkers might have a broader applicability in other forms of synucleinopathies. A study of Umehara and co-authors evaluated the peripheral immune profile in drug-naïve dementia with Lewy bodies (DLB) and reported lower absolute lymphocyte and basophil counts as compared to age-matched healthy controls. Interestingly in this study, higher basophil counts were marginally associated with higher global cognition and significantly associated with milder motor severity (Umehara et al., 2024). Moreover, when assessing Multiple System Atrophy (MSA) patients compared to PD and healthy controls in another study, authors reported significant differences in NLR and monocytes between MSA patients and HCs (higher NLR in MSA).

Monocytes and uric acid (UA) levels were also significantly different between the MSA-P and PD patients (Madetko et al, 2022). In MSA-P patients NLR was significantly higher compared to HC but there was no difference in NLR between patients with MSA-P and PD. It has been proposed that in MSA patients, monocytes, and NLR might serve as potential diagnostic biomarkers, whereas MLR, complement C3 and C4, and immunoglobulin IgG significantly correlate with disease severity (Yuan et al., 2024, Matuse et al., 2020). Interestingly, despite not being a synucleinopathy, the NLR ratio has been reported to serve as a putative marker of peripheral inflammation also in progressive supranuclear palsy (PSP) (Inci et al., 2020).

The NLR Index and absolute or relative measurements of leukocyte subpopulations from peripheral blood represent useful, cost-effective and accessible immunological biomarkers that could be applied to idiopathic or genetic forms of Parkinson’s disease and other neurodegenerative disorders. Our results combined with those from other genetic clinical cohorts could help elucidate the role of neuroinflammation in degenerative disorders and inform future clinical trials especially those targeting patients with defined genetic backgrounds. Regarding implications for therapeutic interventions, novel biomarkers of neuroinflammation in PD and other synucleinopathies (including NLR or other leukocyte-based indexes) could facilitate the identification of early stage participants and enhance a close monitoring for response to novel treatment regimens.

## Data Availability

All data produced in the present study are available upon reasonable request to the authors.

## Aknowledgements

This work was supported by funding from the PPMI study, Parkinson’s Progression Markers Initiative. PPMI - a public-private partnership - is funded by the Michael J. Fox Foundation for Parkinson’s Research and funding partners, including Abbvie, AcureX, Allergan, Aligning Science Across Parkinson’s, Amathus Therapeutics, Avid Radiopharmaceuticals, Bial Biotech, Biohaven, Biogen, BioLegend, BlueRock Therapeutics, Bristol-Myers Squibb, Calico Labs, Celgene, Cerevel, Coave, DaCapo Brainscience, 4D Pharma, Denali, Edmond J. Safra Foundation, Eli Lilly, Gain Therapeutics, GE Healthcare, Genentech, GlaxoSmithKline, Golub Capital, Handl Therapeutics, Insitro, Janssen Neuroscience, Lundbeck, Merck, Meso Scale Discovery, Neurocrine Biosciences, Pfizer, Piramal, Prevail Therapeutics, Roche, Sanofi Genzyme, Servier, Takeda, Teva, UCB, VanquaBio, Verily, Voyager Therapeutics, and Yumanity.

## Notes

### Competing Interest Statement

The authors have declared no competing interest.

### Author Declarations

Data used in the preparation of this article were obtained on 03/27/2023 (08/15/2024 last update) from the PPMI database (http://www.ppmiinfo.org/access-data-specimens/download-data),RRID:SCR 006431. For up-to-date information on the study, visit http://www.ppmi-info.org. Signed informed consent was obtained from all participants recruited. The study was approved by the Scientific Board of all PPMI sites involved (including the Scientific Board of Eginition hospital).

## Reference list

Li F, Weng G, Zhou H, Zhang W, Deng B, Luo Y, Tao X, Deng M, Guo H, Zhu S, Wang Q. The neutrophil-to-lymphocyte ratio, lymphocyte-to-monocyte ratio, and neutrophil-to-high-density-lipoprotein ratio are correlated with the severity of Parkinson’s disease. Front Neurol. 2024 Jan 23;15:1322228. doi: 10.3389/fneur.2024.1322228. eCollection 2024.

Zhang F, Chen B, Ren W, Yan Y, Zheng X, Jin S, Chang Y. Association analysis of dopaminergic degeneration and the neutrophil-to-lymphocyte ratio in Parkinson’s disease. Front Aging Neurosci. 2024 Apr 8;16:1377994. doi: 10.3389/fnagi.2024.1377994. eCollection 2024.

Umehara T, Mimori M, Kokubu T, Ozawa M, Shiraishi T, Sato T, Onda A, Matsuno H, Omoto S, Sengoku R, Murakami H, Oka H, Iguchi Y. Peripheral immune profile in drug-naïve dementia with Lewy bodies. J Neurol. 2024 Apr 6. doi: 10.1007/s00415-024-12336-x.

Novellino F, Donato A, Malara N, Madrigal JL, Donato G. Complete blood cell count-derived ratios can be useful biomarkers for neurological diseases. Int J Immunopathol Pharmacol. (2021) 35:20587384211048264. doi: 10.1177/20587384211048264

Madetko N, Migda B, Alster P, Turski P, Koziorowski D, Friedman A. Platelet-to-lymphocyte ratio and neutrophil-to lymphocyte ratio may reflect differences in PD and MSA-P neuroinflammation patterns. Neurol Neurochir Pol. (2022) 56:148–55. doi: 10.5603/PJNNS.a2022.0014,

Dommershuijsen LJ, Ruiter R, Erler NS, Rizopoulos D, Ikram MA, Ikram MK. Peripheral immune cell numbers and C-reactive protein in Parkinson’s disease: results from a population-based study. J Parkinsons Dis. (2022) 12:667–78. doi: 10.3233/JPD-212914,

Muñoz-Delgado L, Macías-García D, Jesús S, Martín-Rodríguez JF, Labrador-Espinosa M, Jiménez-Jaraba MV, et al. Peripheral immune profile and neutrophil-to-lymphocyte ratio in Parkinson’s disease. Mov Disord. (2021) 36:2426–30. doi: 10.1002/mds.28685,

Liu Z, Fan Q, Wu S, Wan Y, Lei Y. Compared with the monocyte to high-density lipoprotein ratio (MHR) and the neutrophil to lymphocyte ratio (NLR), the neutrophil to high-density lipoprotein ratio (NHR) is more valuable for assessing the inflammatory process in Parkinson’s disease. Lipids Health Dis. (2021) 20:35. doi: 10.1186/s12944-021-01462-4,

Umehara T, Oka H, Nakahara A, Matsuno H, Murakami H. Differential leukocyte count is associated with clinical phenotype in Parkinson’s disease. J Neurol Sci. (2020) 409:116638. doi: 10.1016/j.jns.2019.116638,

Su Y, Shi C, Wang T, Liu C, Yang J, Zhang S, et al. Dysregulation of peripheral monocytes and pro-inflammation of alpha-synuclein in Parkinson’s disease. J Neurol. (2022) 269:6386–94. doi: 10.1007/s00415-022-11258-w,

Muñoz-Delgado L, Macías-García D, Periñán MT, Jesús S, Adarmes-Gómez AD, Bonilla Toribio M, et al. Peripheral inflammatory immune response differs among sporadic and familial Parkinson’s disease. npj Parkinsons Dis. (2023) 9:12. doi: 10.1038/s41531-023-00457-5

Williams GP, Schonhoff AM, Sette A, Lindestam Arlehamn CS. Central and peripheral inflammation: connecting the immune responses of Parkinson’s disease. J Parkinsons Dis. (2022) 12:S129–s136. doi: 10.3233/JPD-223241

Bawa KK, Krance SH, Herrmann N, Cogo-Moreira H, Ouk M, Yu D, et al. A peripheral neutrophil-related inflammatory factor predicts a decline in executive function in mild Alzheimer’s disease. J Neuroinflammation. (2020) 17:84. doi: 10.1186/s12974-020-01750-3

Contaldi E, Magistrelli L, Cosentino M, Marino F, Comi C. Lymphocyte count and neutrophil-to-lymphocyte ratio are associated with mild cognitive impairment in Parkinson’s disease: a single-center longitudinal study. J Clin Med. (2022) 11:5543. doi: 10.3390/jcm11195543,

Matsuse D, Yamasaki R, Maimaitijiang G, Yamaguchi H, Masaki K, Isobe N, et al. Early decrease in intermediate monocytes in peripheral blood is characteristic of multiple system atrophy-cerebellar type. J Neuroimmunol. (2020) 349:577395. doi: 10.1016/j.jneuroim.2020.577395,

Akıl E, Bulut A, Kaplan İ, Özdemir HH, Arslan D, Aluçlu MU. The increase of carcinoembryonic antigen (CEA), high-sensitivity C-reactive protein, and neutrophil/lymphocyte ratio in Parkinson’s disease. Neurol Sci. (2015) 36:423–8. doi: 10.1007/s10072-014-1976-1

Bissacco J, Simonetta C, Mascioli D, Zenuni H, Bovenzi R, Grillo P, Di Giuliano F, Stefani A, Mercuri NB, Schirinzi T. Peripheral immunity changes are associated with neurodegeneration and worse clinical outcome in idiopathic normal pressure hydrocephalus. Eur J Neurol. 2024 Mar;31(3):e16179. doi: 10.1111/ene.16179. Epub 2023 Dec 21.

Hosseini S, Shafiabadi N, Khanzadeh M, Ghaedi A, Ghorbanzadeh R, Azarhomayoun A, Bazrgar A, Pezeshki J, Bazrafshan H, Khanzadeh S. Neutrophil to lymphocyte ratio in parkinson’s disease: a systematic review and meta-analysis. BMC Neurol. 2023 Sep 21;23(1):333.

Kim R, Kang N, Byun K, Park K, Jun JS. Prognostic significance of peripheral neutrophils and lymphocytes in early untreated Parkinson’s disease: an 8-year follow-up study. J Neurol Neurosurg Psychiatry. 2023 Dec;94(12):1040–1046.

Grillo P, Sancesario GM, Bovenzi R, Zenuni H, Bissacco J, Mascioli D, Simonetta C, Forti P, Degoli GR, Pieri M, Chiurchiù V, Stefani A, Mercuri NB, Schirinzi T. Neutrophil-to-lymphocyte ratio and lymphocyte count reflect alterations in central neurodegeneration-associated proteins and clinical severity in Parkinson Disease patients. Parkinsonism Relat Disord. 2023 Jul;112:105480. doi: 10.1016/j.parkreldis.2023.10548

Muñoz-Delgado L, Labrador-Espinosa MÁ, Macías-García D, Jesús S, Benítez Zamora B, Fernández-Rodríguez P, Adarmes-Gómez AD, Reina Castillo MI, Castro-Labrador S, Silva-Rodríguez J, Carrillo F, García Solís D, Grothe MJ, Mir P. Peripheral Inflammation Is Associated with Dopaminergic Degeneration in Parkinson’s Disease. Mov Disord. 2023 May;38(5):755–763. doi: 10.1002/mds.29369.0

Wang LX, Liu C, Shao YQ, Jin H, Mao CJ, Chen J. Peripheral blood inflammatory cytokines are associated with rapid eye movement sleep behavior disorder in Parkinson’s disease. Neurosci Lett. 2022 Jun 21;782:136692. doi: 10.1016/j.neulet.2022.136692.

Kara SP, Altunan B, Unal A. Investigation of the peripheral inflammation (neutrophil-lymphocyte ratio) in two neurodegenerative diseases of the central nervous system. Neurol Sci. 2022 Mar;43(3):1799–1807. doi: 10.1007/s10072-021-05507-5.

Jensen MP, Jacobs BM, Dobson R, Bandres-Ciga S, Blauwendraat C, Schrag A, Noyce AJ; International Parkinson’s Disease Genomics Consortium (IPDGC). Lower Lymphocyte Count is Associated With Increased Risk of Parkinson’s Disease. Ann Neurol. 2021 Apr;89(4):803–812. doi: 10.1002/ana.2603

Inci I, Kusbeci OY, Eskut N. The neutrophil-to-lymphocyte ratio as a marker of peripheral inflammation in progressive supranuclear palsy: a retrospective study. Neurol Sci. 2020 May;41(5):1233–1237. doi: 10.1007/s10072-019-04208-4. Epub 2020 Jan 4.

Yuan X, Wan L, Chen Z, Long Z, Chen D, Liu P, Fu Y, Zhu S, Peng L, Qiu R, Tang B, Jiang H. Peripheral Inflammatory and Immune Landscape in Multiple System Atrophy: A Cross-Sectional Study. Mov Disord. 2024 Feb;39(2):391–399.

Papadimitriou D, Antonelou R, Miligkos M, Maniati M, Papagiannakis N, Bostantjopoulou S, Leonardos A, Koros C, Simitsi A, Papageorgiou SG, Kapaki E, Alcalay RN, Papadimitriou A, Athanassiadou A, Stamelou M, Stefanis L. Motor and Nonmotor Features of Carriers of the p.A53T Alpha-Synuclein Mutation: A Longitudinal Study. Mov Disord. 2016 Aug;31(8):1226–30. doi: 10.1002/mds.26615.

Koros C, Stamelou M, Simitsi A, Beratis I, Papadimitriou D, Papagiannakis N, Fragkiadaki S, Kontaxopoulou D, Papageorgiou SG, Stefanis L. Selective cognitive impairment and hyposmia in p.A53T SNCA PD vs typical PD. Neurology. 2018 Mar 6;90(10):e864–e869. doi: 10.1212/WNL.0000000000005063.

Hiraga K, Hattori M, Satake Y, Tamakoshi D, Fukushima T, Uematsu T, Tsuboi T, Sato M, Yokoi K, Suzuki K, Arahata Y, Washimi Y, Hori A, Yamamoto M, Shimizu H, Wakai M, Tatebe H, Tokuda T, Nakamura A, Niida S, Katsuno M. Plasma biomarkers of neurodegeneration in patients and high risk subjects with Lewy body disease. NPJ Parkinsons Dis. 2024 Jul 31;10(1):135. doi: 10.1038/s41531-024-00745-8.

Chopra A, Outeiro TF. Aggregation and beyond: alpha-synuclein-based biomarkers in synucleinopathies. Brain. 2024 Jan 4;147(1):81–90. doi: 10.1093/brain/awad260.

Park Y, Kc N, Paneque A, Cole PD. Tau, Glial Fibrillary Acidic Protein, and Neurofilament Light Chain as Brain Protein Biomarkers in Cerebrospinal Fluid and Blood for Diagnosis of Neurobiological Diseases. Int J Mol Sci. 2024 Jun 7;25(12):6295. doi: 10.3390/ijms25126295.

Lindestam Arlehamn CS, Dhanwani R, Pham J, Kuan R, Frazier A, Rezende Dutra J, Phillips E, Mallal S, Roederer M, Marder KS, Amara AW, Standaert DG, Goldman JG, Litvan I, Peters B, Sulzer D, Sette A. α-Synuclein-specific T cell reactivity is associated with preclinical and early Parkinson’s disease. Nat Commun. 2020 Apr 20;11(1):1875. doi: 10.1038/s41467-020-15626-w.

Castelo Rueda MP, Raftopoulou A, Gögele M, Borsche M, Emmert D, Fuchsberger C, Hantikainen EM, Vukovic V, Klein C, Pramstaller PP, Pichler I, Hicks AA. Frequency of Heterozygous Parkin (PRKN) Variants and Penetrance of Parkinson’s Disease Risk Markers in the Population-Based CHRIS Cohort. Front Neurol. 2021 Aug 9;12:706145. doi: 10.3389/fneur.2021.706145. eCollection 2021.

Chen Z, Chen S, Liu J. The role of T cells in the pathogenesis of Parkinson’s disease. Prog Neurobiol. 2018 Oct;169:1–23. doi: 10.1016/j.pneurobio.2018.08.002.

